# Herd immunity thresholds for SARS-CoV-2 estimated from unfolding epidemics

**DOI:** 10.1101/2020.07.23.20160762

**Authors:** Ricardo Aguas, Guilherme Gonçalves, Marcelo U. Ferreira, M. Gabriela M. Gomes

## Abstract

Variation in individual susceptibility or frequency of exposure to infection accelerates the rate at which populations acquire immunity by natural infection. Individuals that are more susceptible or more frequently exposed tend to be infected earlier and hence more quickly selected out of the susceptible pool, decelerating the incidence of new infections as the epidemic progresses. Eventually, susceptible numbers become low enough to prevent epidemic growth or, in other words, the herd immunity threshold (HIT) is reached. We have recently proposed a method whereby mathematical models, with gamma distributions of susceptibility or exposure to SARS-CoV-2, are fitted to epidemic curves to estimate coefficients of individual variation among epidemiological parameters of interest. In the initial study we estimated HIT around 25-29% for the original Wuhan virus in England and Scotland. Here we explore the limits of applicability of the method using Spain and Portugal as case studies. Results are robust and consistent with England and Scotland, in the case of Spain, but fail in Portugal due to particularities of the dataset. We describe failures, identify their causes, and propose methodological extensions.

## Introduction

Selection acting on unmeasured individual variation is a well-known source of bias in the analysis of populations. It has been shown to affect measured rates of mortality (Keyfitz and Littman; Vaupel et al 1979; Vaupel and Yashin 1985), the survival of endangered species (Kendall and Fox 2002; Jenouvrier et al 2018), the scope of neutral theories of biodiversity and molecular evolution (Steiner and Tuljapurkar 2012, Gomes et al 2019), the measured risk of diseases whether non-communicable (Aalen et al 2015; Stensrud and Valberg 2017) or infectious (Anderson et al 1986; Dwyer et al 1997; Smith et al 2005; Bellan et al 2015; Gomes et al 2019; Corder et al 2020; Montalbán et al 2020), and the efficacy of interventions such as vaccines (Halloran et al 1996; O’Hagan et al 2012; Gomes et al 2014; Gomes et al 2016; Langwig et al 2017) or symbionts (Pessoa et al 2016; King et al 2018). Building on this knowledge, we previously addressed how selection on individual variation might affect the course of the coronavirus disease (COVID-19) pandemic (Gomes et al 2022).

COVID-19 is an infectious respiratory disease caused by a virus (severe acute respiratory syndrome coronavirus 2 [SARS-CoV-2]), which was first identified in China in late 2019 and has since spread worldwide leading to considerable human suffering and social disruption. European and American continents have been the most affected, with 0.16% and 0.20% of the respective total populations having died as of the 15 July 2021 (WHO 2021). Here we analyse series of daily deaths attributed to COVID-19 in Spain and Portugal (Iberian Peninsula) to study how individual variation in susceptibility and exposure to a respiratory virus affects its epidemic trajectory. Besides adding to the compendium of neglected effects of selection in population dynamics we hope to stimulate a new approach to study epidemic dynamics.

The essence to the approach is that individual variation in susceptibility or exposure (connectivity) accelerates the acquisition of immunity in populations. More susceptible and more connected individuals have a higher propensity to be infected and thus are likely to become immune earlier. Due to this selective immunization by natural infection, heterogeneous populations acquire herd immunity more efficiently than suggested by models that do not fully account for these types of variation. Here we integrate a continuous distribution of susceptibility or connectivity in an otherwise basic COVID-19 epidemiological model, which necessarily accounts for non-pharmaceutical intervention effects, and generate three types of results. First, at national levels the herd immunity threshold by natural infection declines from around 70% to 20-30%. This is newly reported here for Spain and in agreement with recent estimates for England and Scotland (Gomes et al 2022). Second, these inferences can be made relatively early in the pandemic, such as between first and second waves, provided the first wave is sufficiently large and spatially synchronous. Third, we include a selection of results for Portugal to illustrate how the inferential procedure degenerates when national data do not meet certain conditions.

### Individual variation in SARS-CoV-2 transmission

SARS-CoV-2 is transmitted primarily by respiratory droplets and modelled as a susceptible-exposed-infectious-recovered (SEIR) process.

#### Variation in susceptibility to infection

Individual variation in susceptibility is integrated as a continuously distributed factor that multiplies the force of infection upon individuals (Diekmann et al 1990) in the form of an infinite system of ordinary differential equations (ODEs):

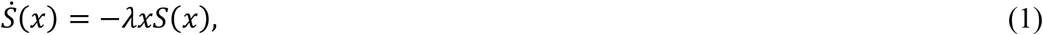

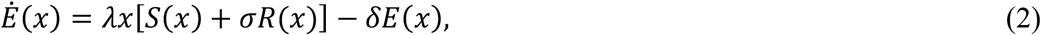

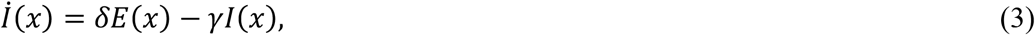

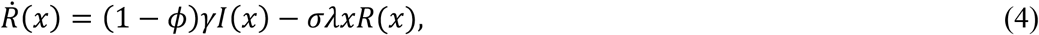

where *S*(*x*) is the density of individuals with susceptibility *x, E*(*x*) and *I*(*x*) are the densities of individuals who originally had susceptibility *x* and became exposed and infectious, while *R*(*x*) represents those who have recovered and have their susceptibility reduced to a reinfection factor *σ* due to acquired immunity. Parameter *δ* is the rate of progression from exposed to a period of maximal infectiousness (= 1/5.5 per day [McAloon et al. 2020; Lauer et al. 2020]), *γ* is the rate of recovery from maximal infectiousness (= 1/4 per day [Nishiura et al. 2020; Li et al. 2020]), *ϕ* is the proportion of individuals who die as a result of infection (= 0.008 [Pastor-Barriuso et al. 2021]), and:

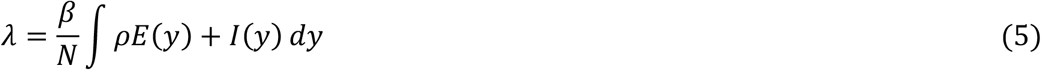

is the average force of infection upon susceptible individuals in a population of approximately constant size *N* and transmission coefficient *β*. Standardizing so that susceptibility distributions have mean ∫ *xg*(*x*) *dx* = 1, given a probability density function *g*(*x*), the basic reproduction number, defined as the expected number of secondary infections generated by an infected individual in a population that has no specific immunity to the virus (Diekmann et al 1990), is:

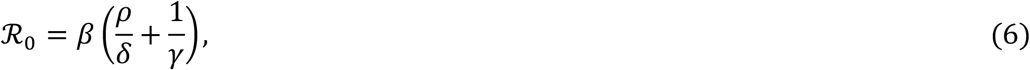

where *ρ* is a factor measuring the infectiousness of individuals in compartment *E* in relation to those in *I* (= 0.5 [Wei et al. 2020; To et al. 2020; Arons et al. 2020; He et al. 2020]). The coefficient of variation (CV) in individual susceptibility 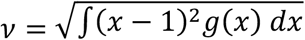 is also treated as a parameter.

The basic reproduction number *ℛ*_0_ is a theoretical framework. It is usually estimated from the initial growth in case numbers. However, as the virus spreads through the population, infected and immune individuals accumulate, reducing the availability of susceptible hosts. As a result, growth in case numbers deviates from being a direct indication of *ℛ*_0_ but rather of a so-called effective reproduction number *ℛ*_*eff*_.

When susceptibility is given by a gamma distribution and acquired immunity is totally protective (*σ* = 0), the effective reproduction number is:

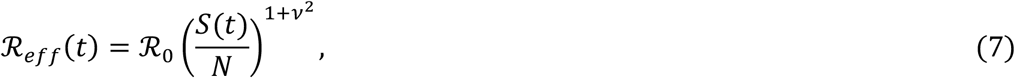

where *S*(*t*) = ∫ *S*(*x, t*) *dx* is the total number of susceptible individuals at time *t*, and (Equations 1-4) reduce exactly to a finite system of ODEs:

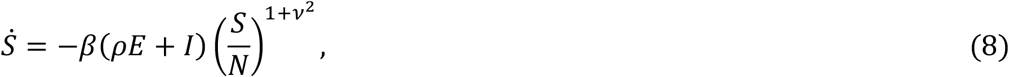

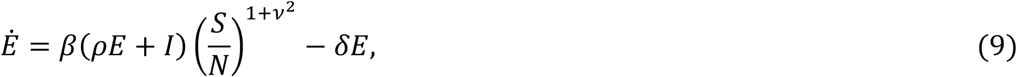

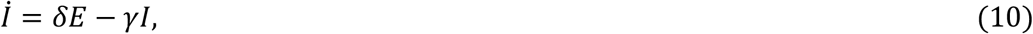

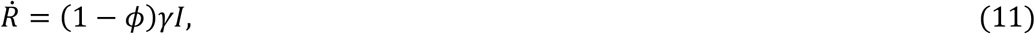

where *S, E, I* and *R* are the total numbers of susceptible, exposed, infectious and recovered individuals, respectively (Novozhilov 2008; Montalbán et al. 2020).

#### Variation in connectivity

In a directly transmitted infectious disease, such as COVID-19, variation in exposure to infection is governed primarily by patterns of connectivity among individuals. Here we incorporate this in the system (Equations 1-4) under the assumption that individuals mix at random (Pastor-Satorras and Vespignani 2001; Miller et al. 2012), while in Supplementary Information we conduct some sensitivity analyses to this assumption. Under random mixing and heterogeneous connectivity, the force of infection is written as:

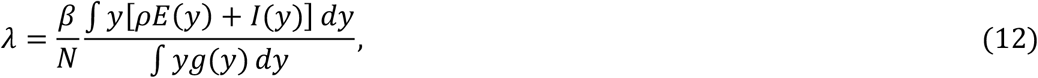

and the basic reproduction number is:

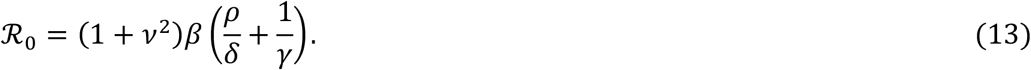

In this setup, when connectivity is given by a gamma distribution and acquired immunity is totally protective, the effective reproduction number is approximated by:

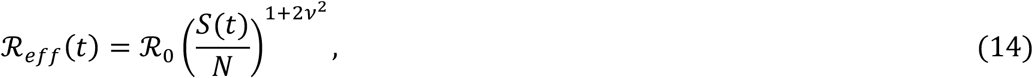

and (Equations 1-4) reduce approximately to the finite system of ODEs:

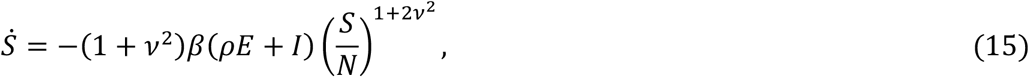

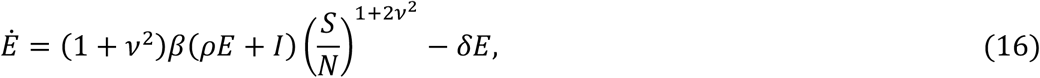

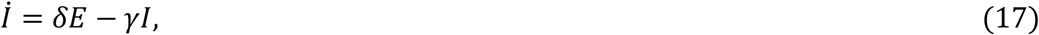

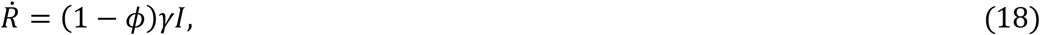

where *S, E, I* and *R* are the total numbers of susceptible, exposed, infectious and recovered individuals, respectively (Montalbán et al. 2020).

We have combined the basic models described by the reduced systems in Equations 8-11 and Equations 15-18 with non-pharmaceutical interventions (NPIs) to produce COVID-19 transmission models. We then estimate the relevant parameters for those models by fitting to data series of daily deaths from each of the study countries.

### Non-pharmaceutical interventions and other transmissibility modifiers

NPIs designed to control transmission typically reduce *β* and hence *ℛ*_0_. Denoting the time-dependent reproduction number when control measures are in place by *ℛ*_*c*_(*t*), the modified effective reproduction number is obtained by replacing *ℛ*_0_ with *ℛ*_*c*_(*t*) in (Equation 7) and (Equation 14) as appropriate. For the estimation of *ℛ*_*c*_(*t*) we introduce flexible transmissibility profiles *c*(*t*) as illustrated in Figure 1.

**Figure 1:**
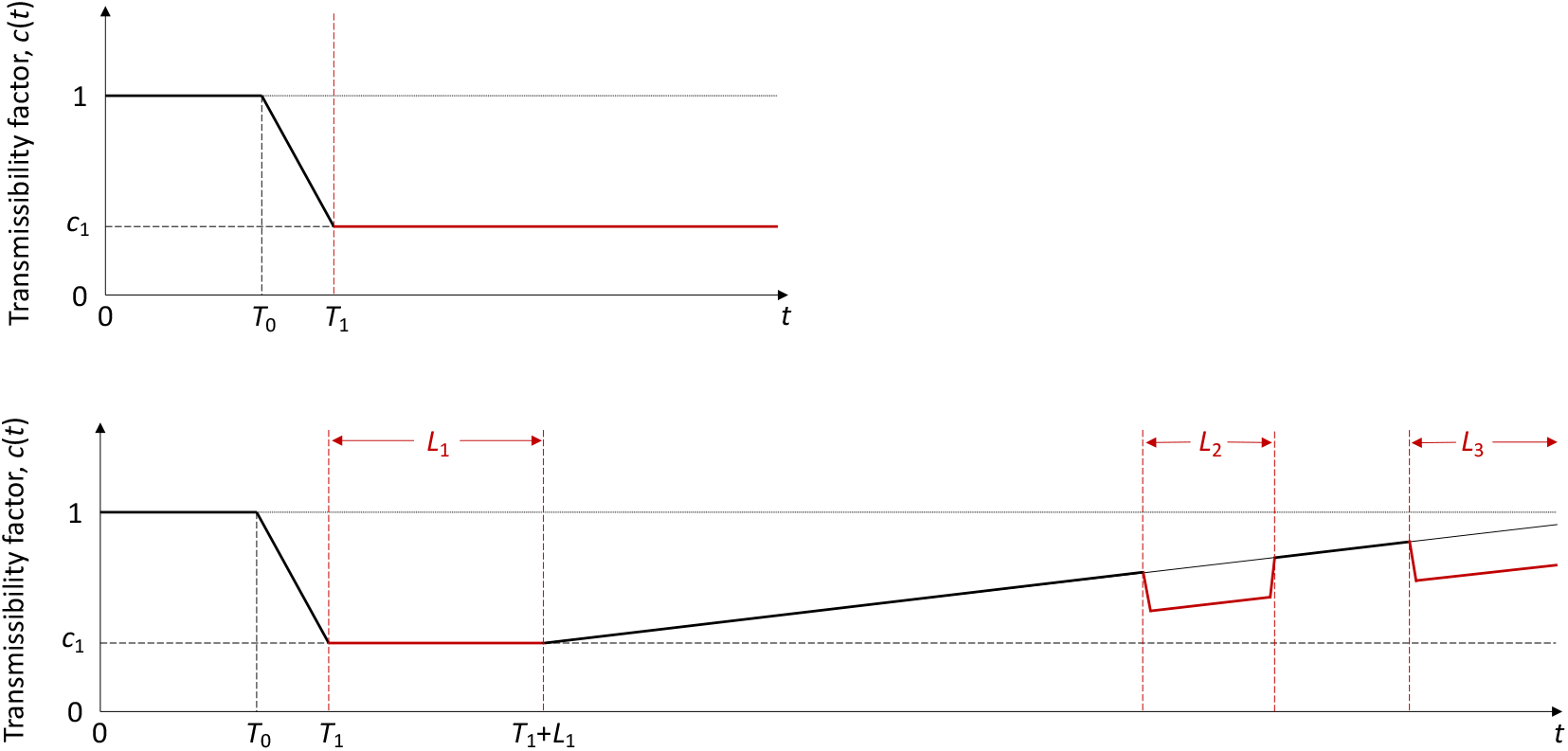
Transmissibility profile. Schematic illustration of factor *c*(*t*) representing the combined effects of NPIs, seasonality and viral evolution on the reproduction number *ℛ*_*c*_(*t*). *T*_0_ is the time when *ℛ*_*c*_ begins to decrease due to behavioural change or seasonality (estimated); *T*_1_ (> *T*_0_) is the day first lockdown begins (informed by data); *c*_1_ ≤ 1 is the average *c*(*t*) achieved during the first lockdown; *L*_1_, *L*_2_ and *L*_3_, denote the length in days of the successive periods of strictest NPI measures (*L*_1_ being the first lockdown). These profiles are adopted in fits until: 1 July 2020 (top); 1 March 2021 (bottom).

#### One-wave transmissibility profile

When the model is applied to the first pandemic wave only (until 1 July 2020) we use the top profile in Figure 1. *T*_0_ is the time when *ℛ*_0_ begins to decrease due to behavioural changes or seasonality; *T*_1_ (> *T*_0_) is the day first lockdown begins (transmission is allowed to decrease between *T*_0_ and *T*_1_); *c*_1_ ≤ 1 is the average *c*(*t*) from the beginning of the first lockdown onwards (14 March in Spain, 19 March in Portugal). Mathematically this is constructed as:

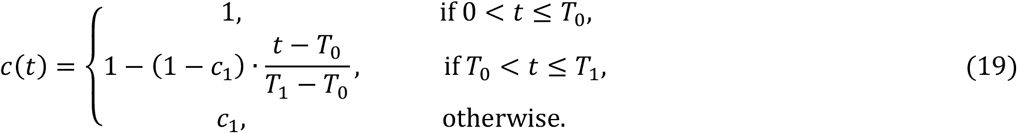

#### Two-wave transmissibility profile

Applying the model over longer periods which capture multiple waves and multiple lockdowns requires additional features on the transmissibility profile. Denoting by *L*_1_ the duration of the first lockdown (29 days in Spain, 44 days in Portugal), we allow restrictions to be progressively relaxed at the end of this period by letting transmission begin a linear increase such that *c*(*t*) reaches 1 in *T*_2_ days, which may or may not be within the range of the study. Changes in other factors that affect transmission (such as seasonality or viral evolution) are inseparable from contact changes in this framework and are also accounted for by *c*(*t*). Mathematically this is constructed as:

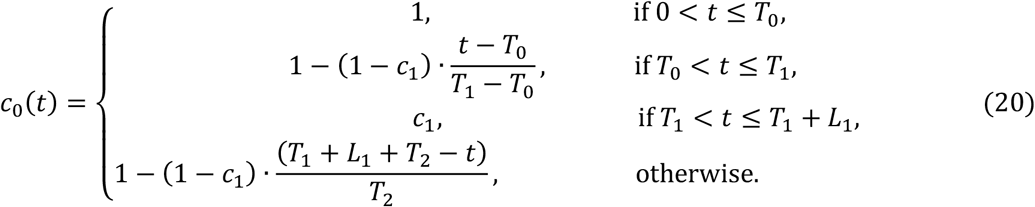

Second and third lockdowns in the autumn and winter season are implemented as a further reduction in transmission (by factors *c*_2_ and *c*_3_, respectively) over the stipulated time periods (*L*_2_ and *L*_3_ in the bottom panel of Figure 1):

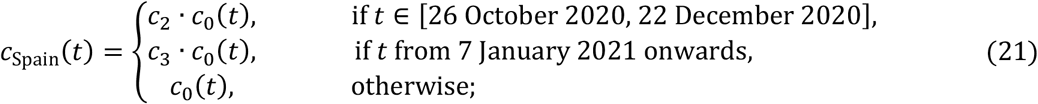

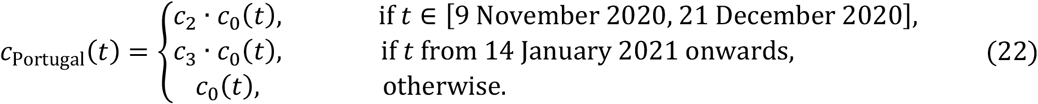

In Spain, second and third lockdowns were effectively a single intervention, moderately interrupted by a short relaxation over Christmas, and hence we assume *c*_2_ = *c*_3_ in this country. This is contrasted by Portugal where the third lockdown was much stricter than the second and estimated independently.

### Herd immunity thresholds

Individual variation in risk of acquiring infection is under selection by the force of infection, whether individual differences are due to biological susceptibility, exposure, or both. The most susceptible or exposed individuals are selectively removed from the susceptible pool as they become infected and eventually recover with immunity (some die), resulting in decelerated epidemic growth and accelerated acquisition of immunity in the population. The herd immunity threshold (HIT) defines the percentage of the population that needs to be immune to reverse epidemic growth and prevent future waves. In the absence of NPIs or other transmissibility modifiers, if individual susceptibility or connectivity is gamma-distributed and mixing is random, basic HIT curves (*ℋ*) can be derived analytically (Montalbán et al 2020) from the model systems (Equations 1-4, with the respective forces of infections). In the case of variation in susceptibility to infection we obtain

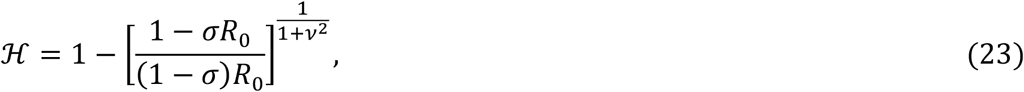

while variable connectivity results in a different exponent:

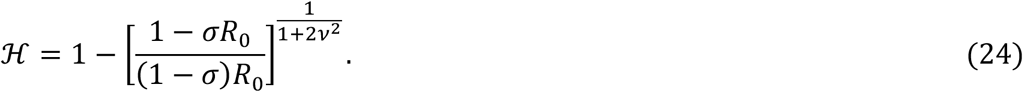

In less straightforward cases, such as when the characteristics follow a distribution other than gamma (Gomes et al. 2022), when mixing is not random (Supplementary Information), when both distributions in susceptibility and connectivity are considered, or when contact networks are rewired as result of NPIs or otherwise, *ℋ* can be obtained numerically.

In the absence of reinfection (*σ* = 0), both (Equation 23) and (Equation 24) convey substantial declines in HIT as individual variation increases (Figure 2, Gomes et al. 2022 and Montalbán et al. 2020), most strikingly over relatively low CV (from *ν* = 0 up to 1 or 2). For concreteness, when *ℛ*_0_ = 3, *ℋ* = 67% for *ν* = 0, while *ν* = 1 brings *ℋ* down to 42% for heterogeneous susceptibility and 30% for heterogeneous connectivity, and *ν* = 2 brings *ℋ* further down to 20% and 11%, respectively. Accounting for reinfection (*σ* > 0) might moderate these reductions. In any case, such differences in HIT are an indication of strong sensitivity of the epidemic dynamics to the parameter *ν* over a range that appears realistic (Gomes et al. 2022).

**Figure 2:**
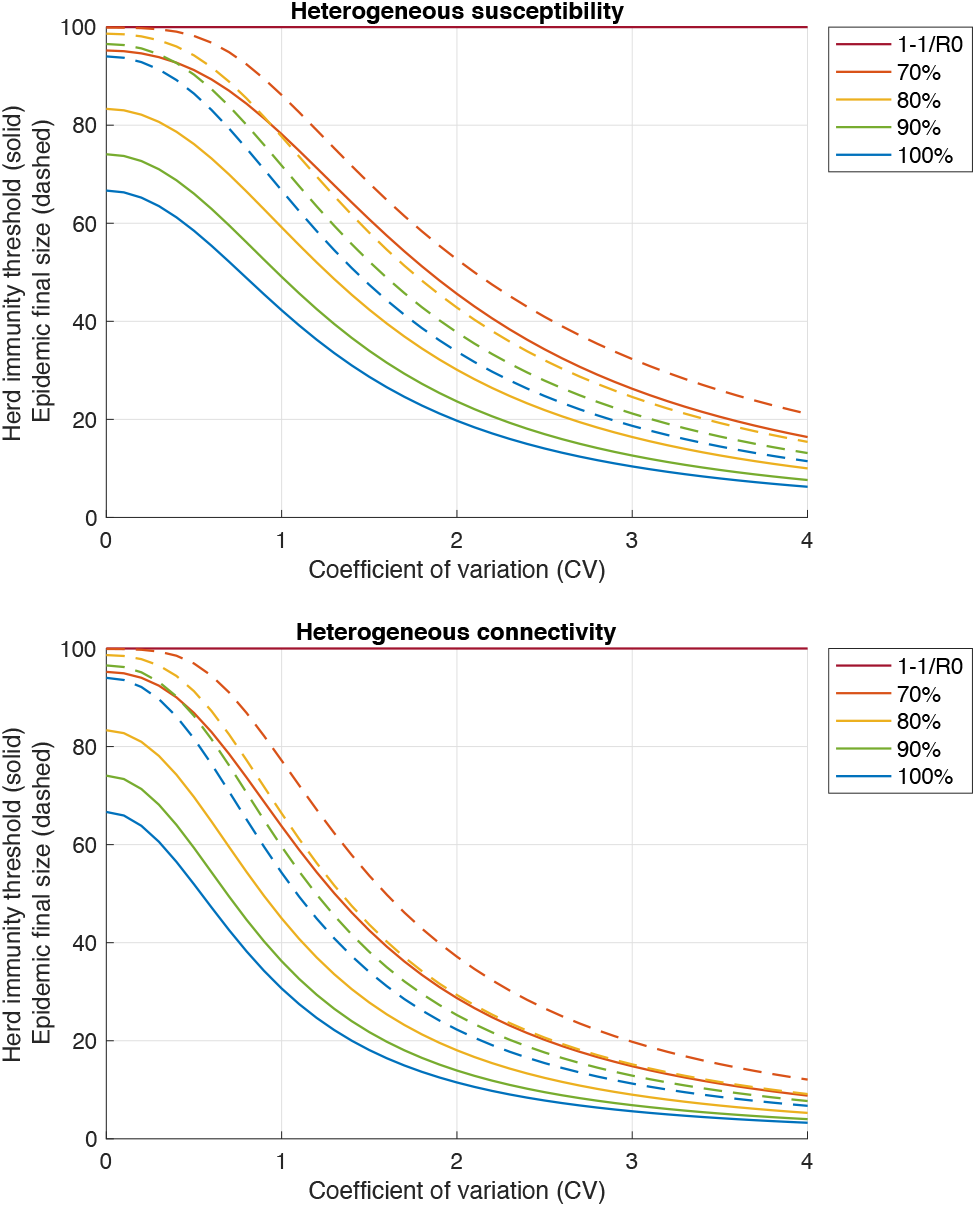
Herd immunity threshold and epidemic final size. Herd immunity thresholds (solid curves) are calculated according to (Equation 23) for heterogeneous susceptibility and (Equation 24) for heterogeneous connectivity, assuming *ℛ*_0_ = 3 for concreteness. Final sizes of the corresponding unmitigated epidemics are also shown (dashed). Curves are generated for different values of the efficacy of immunity conferred by natural infection (1 − *σ*) as displayed in the legend: 100% (blue); 90% (green); 80% (yellow); 70% (orange); 67% (corresponding to 1 − 1/*ℛ*_0_ in this case; red).

We emphasise, nevertheless, that *ℋ* is a theoretical framework to the extent that *ℛ*_0_ is a theoretical framework. It cannot be measured directly when epidemic trajectories are affected by interventions, but it can be inferred indirectly from epidemiological data. By construction, *ℋ* changes if the parameters that determine its value change. Most notably, natural changes in *ℛ*_0_ through time, which can happen due to seasonal forces or viral evolution, transfer to *ℋ* according to (Equation 23), (Equation 24), or even their homogeneity equivalent 1 − 1/*ℛ*_0_. As a result, the percentage of the population immune required to prevent sustained epidemic growth may deviate from the initial *ℋ*. Notwithstanding, a model with lower *ℋ* results in smaller epidemics than a model with higher *ℋ*, all non-basic processes being the same.

The phenomenon of variation and selection which accounts for lower HIT was widely explained around mid-2020 (Hartnett 2020) and generated broad public interest in the context of COVID-19. By early 2021, vaccines had become available, and a competing belief emerged to imply that the HIT might be unachievable for COVID-19 (Aschwanden 2021). While Hartnett (2020) writes about using the basic *ℋ* to assess pandemic potential, stressing how that is weakened by variation and selection by natural infection, Aschwanden (2021) focuses on the imperfect nature of both vaccine induced and natural immunity to endorse that reinfection may become frequent enough to make herd immunity unachievable. In the light of the theory presented here – specifically (Equation 23) and (Equation 24) – these views are orthogonal and do not contradict each other.

### Data

We use publicly available epidemiological data from the coronavirus dashboards for Spain [https://cnecovid.isciii.es/covid19] and Portugal [https://covid19.min-saude.pt/ponto-de-situacao-atual-em-portugal] to fit the models and estimate parameters of interest. Namely, we fit model reconstructed mortality timeseries assuming a fixed infection fatality ratio (IFR) to datasets containing daily deaths, 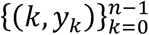, where *k* = 0 is the day when the cumulative moving average of death numbers exceed 5 · 10^−7^ of the population (7 March in Spain, 19 March 2020 in Portugal).

Model fits were carried out to the raw series of daily deaths until the 1 July 2020 in the first instance (to cover the first wave of the epidemic in the study countries), and until the 1 March 2021 in an extended analysis (as a compromise between having a series sufficiently long to capture much of the second wave and not so long that it would be affected by vaccination and require the vaccine to be modelled). We defined the initial conditions as:

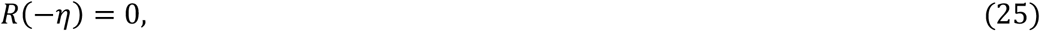

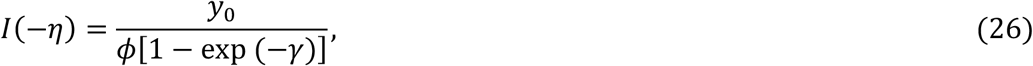

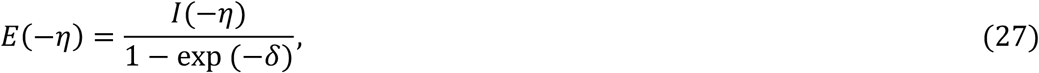

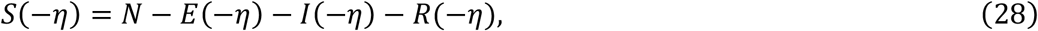

where *η* is the excess duration of a fatal infection relative to non-fatal, *y*_0_ is the number of deaths in the first day of the study, and the population size *N* was obtained from the most recent respective censuses (approx. 46.94 million in Spain, 10.28 million in Portugal, 3.57 million in the North Region of Portugal, 3.66 million in Lisbon and Tagus Valley Region of Portugal).

### Model fitting and parameter estimating

We assumed that reinfection was negligible throughout the study period. A study conducted in England (Hall et al. 2021), between June 2020 and January 2021, concluded that previous SARS-CoV-2 infection induced 84% effective immunity to future infections. In (Gomes et al. 2122) we fit models to daily COVID-19 deaths in England and Scotland, assuming no reinfection (i.e. 100% effective immunity) or 90% effective immunity, and found it to have no significant effect on projected model trajectories. Basically, the fit readjusts the parameters when reinfection is added to the model in such a way that the HIT remains similar.

Parameter estimation was performed with the software MATLAB by employing a multi-start local optimization approach followed by Markov chain Monte Carlo (MCMC) posterior distribution sampling, using the PESTO (Parameter EStimation Toolbox) package (Stapor et al. 2018). We assumed the daily number of SARS-CoV-2 infections to be Poisson distributed.

We approximate the dynamics of COVID-19 deaths by estimating the set of parameters *θ* that maximises the log-likelihood (LL) of observing the daily numbers of reported deaths *Y*:

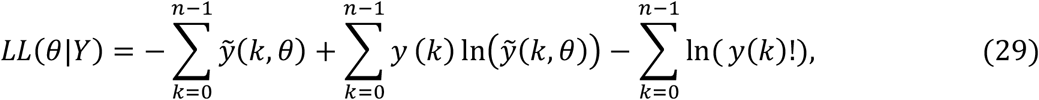

where 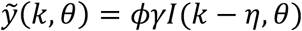 are the simulated model output numbers of COVID-19 deaths at day *k* for the set of parameters *θ*, 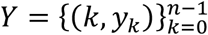 are the numbers of daily reported deaths, and *n* is the total number of days included in the analysis.

### Fitting models to one pandemic wave

The models exploring heterogeneity in susceptibility (Equations 8-11) and connectivity (Equations 15-18) both with transmissibility profile as in (Equation 19), were fit to COVID-19 daily reported deaths in Spain and Portugal recorded until 1 July 2020. A homogeneous version obtained by setting *ν* = 0 in either model was also fitted. Results for Spain are shown in Table 1 and Figure 3, and for Portugal in Table 2 and Figure 4.

**Table 1:** Model parameters for Spain (one wave). Estimated by Bayesian inference based on daily deaths until 1 July 2020. Model selection based on maximum log-likelihood (LL) and Akaike information criterion (AIC). Best fitting models have lower AIC scores (best in red, second best in blue). Herd immunity threshold (*ℋ*) derived from estimated *ℛ*_0_ and CV (*ν*).

|  | Heterogeneous susceptibility |  | Heterogeneous connectivity |  | Homogeneous |  |
| --- | --- | --- | --- | --- | --- | --- |
|  | Median | 95% CI | Median | 95% CI | Median | 95% CI |
| *Estimated parameters* |  |  |  |  |  |  |
| $T_0$ | 5.44 | [3.52, 6.07] | 5.40 | [3.45, 6.07] | 5.48 | [3.73, 6.04] |
| $c_1$ | 0.24 | [0.21, 0.27] | 0.24 | [0.21, 0.28] | 0.18 | [0.18, 0.19] |
| $\eta$ | 16* | [15.53, 16.47] | 16* | [15.52, 16.48] | 16* | [15.52, 16.47] |
| $\mathcal{R}_0$ | 3.59 | [3.51, 3.79] | 3.59 | [3.51, 3.80] | 3.53 | [3.47, 3.71] |
| $\nu$ | 1.98 | [1.33, 2.44] | 1.41 | [0.97, 1.74] | 0 | - |

| *Derived from estimated parameters* |  |  |  |  |  |  |
| --- | --- | --- | --- | --- | --- | --- |
| $\mathcal{H}$ | 23% | [17%, 38%] | 23% | [16%, 37%] | 72% | [71%, 73%] |
| *Model selection* |  |  |  |  |  |  |
| LL | -503.78 |  | -503.50 |  | -518.33 |  |
| AIC | **1017.60** |  | **1017.00** |  | **1044.70** |  |

\* Rounded median resulting in integer number of days.

**Table 2:** Model parameters for Portugal (one wave). Estimated by Bayesian inference based on daily deaths until 1 July 2020. Model selection based on maximum log-likelihood (LL) and Akaike information criterion (AIC). Best fitting models have lower AIC scores (best in red, second best in blue). Herd immunity threshold (*ℋ*) derived from estimated *ℛ*_0_ and CV (*ν*).

|  | Heterogeneous susceptibility |  | Heterogeneous connectivity |  | Homogeneous |  |
| --- | --- | --- | --- | --- | --- | --- |
|  | Median | 95% CI | Median | 95% CI | Median | 95% CI |
| *Estimated parameters* |  |  |  |  |  |  |
| $T_0$ | 0.05 | [0.00, 0.10] | 0.05 | [0.00, 0.10] | 0.05 | [0.00, 0.10] |
| $c_1$ | 0.31 | [0.26, 0.36] | 0.32 | [0.26, 0.39] | 0.31 | [0.26, 0.35] |
| $\eta$ | 20* | [14.75, 25.72] | 20* | [13.27, 25.94] | 21* | [15.56, 26.04] |
| $\mathcal{R}_0$ | 2.62 | [2.29, 3.21] | 2.66 | [2.27, 3.39] | 2.61 | [2.28, 3.15] |
| $\nu$ | 0.31 | [0.01, 3.24] | 0.38 | [0.01, 3.04] | 0 | - |
| *Derived from estimated parameters* |  |  |  |  |  |  |
| $\mathcal{H}$ | 58% | [7%, 69%] | 53% | [4%, 70%] | 62% | [56%, 68%] |
| *Model selection* |  |  |  |  |  |  |
| LL | -287.10 |  | -287.14 |  | -287.34 |  |
| AIC | **584.20** |  | **584.28** |  | **582.67** |  |

\* Rounded median resulting in integer number of days.

**Figure 3:**
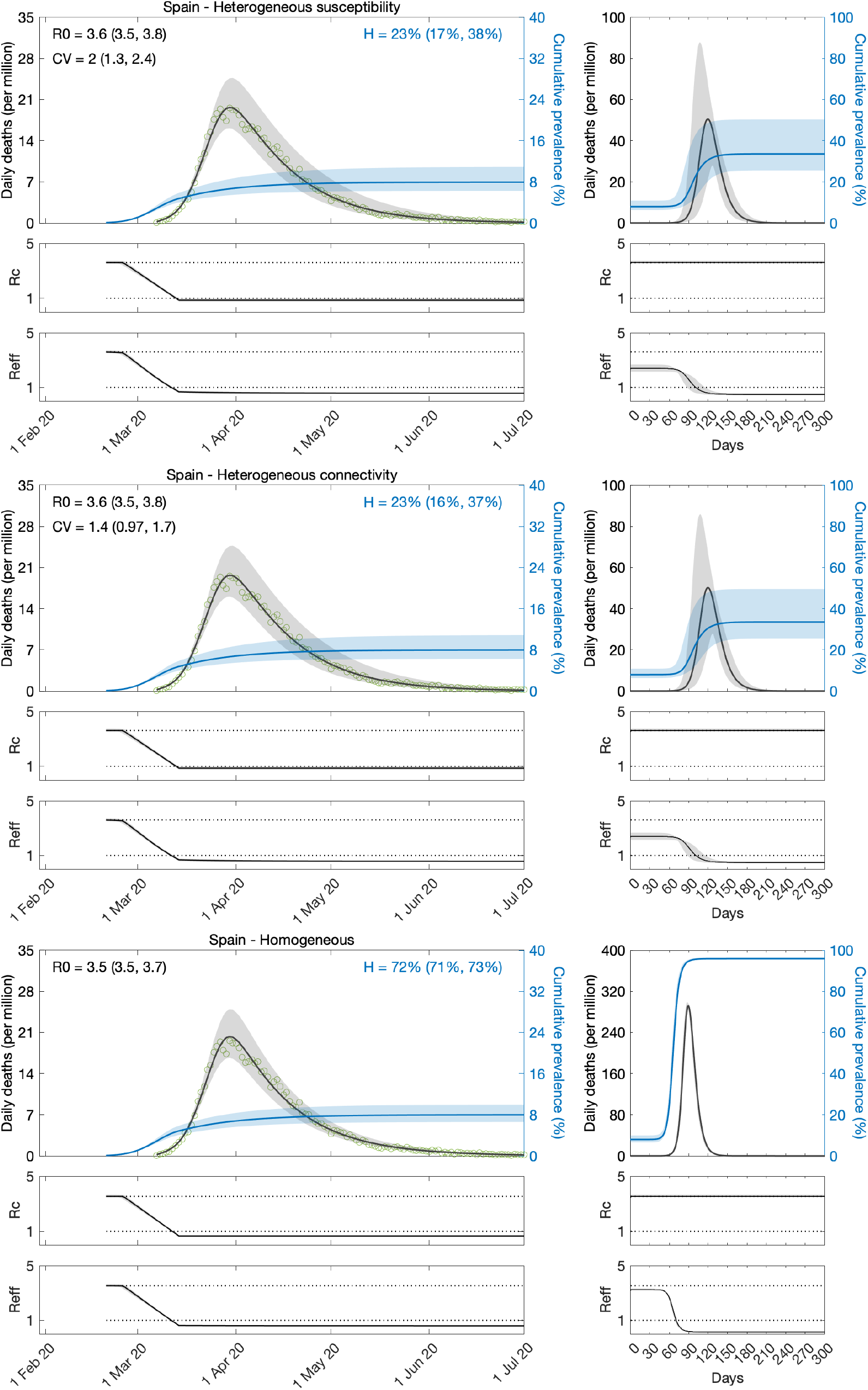
Estimating SARS-CoV-2 transmission in Spain by fitting one wave of COVID-19 deaths. Variation in susceptibility (top panels); variation in connectivity (middle panels); and homogeneous model (bottom panels). Susceptibility or connectivity factors implemented as gamma distributions. Controlled (*ℛ*_*c*_) and effective (*ℛ*_*eff*_) reproduction numbers are displayed on shallow panels underneath the main plots. Basic reproduction number, coefficients of variation and transmissibility profile parameters estimated by Bayesian inference as described in Methods (estimates in Table 1). Curves represent model reconstructions from the median posterior parameter estimates. Shades represent 95% credible intervals from 100,000 posterior samples.

**Figure 4:**
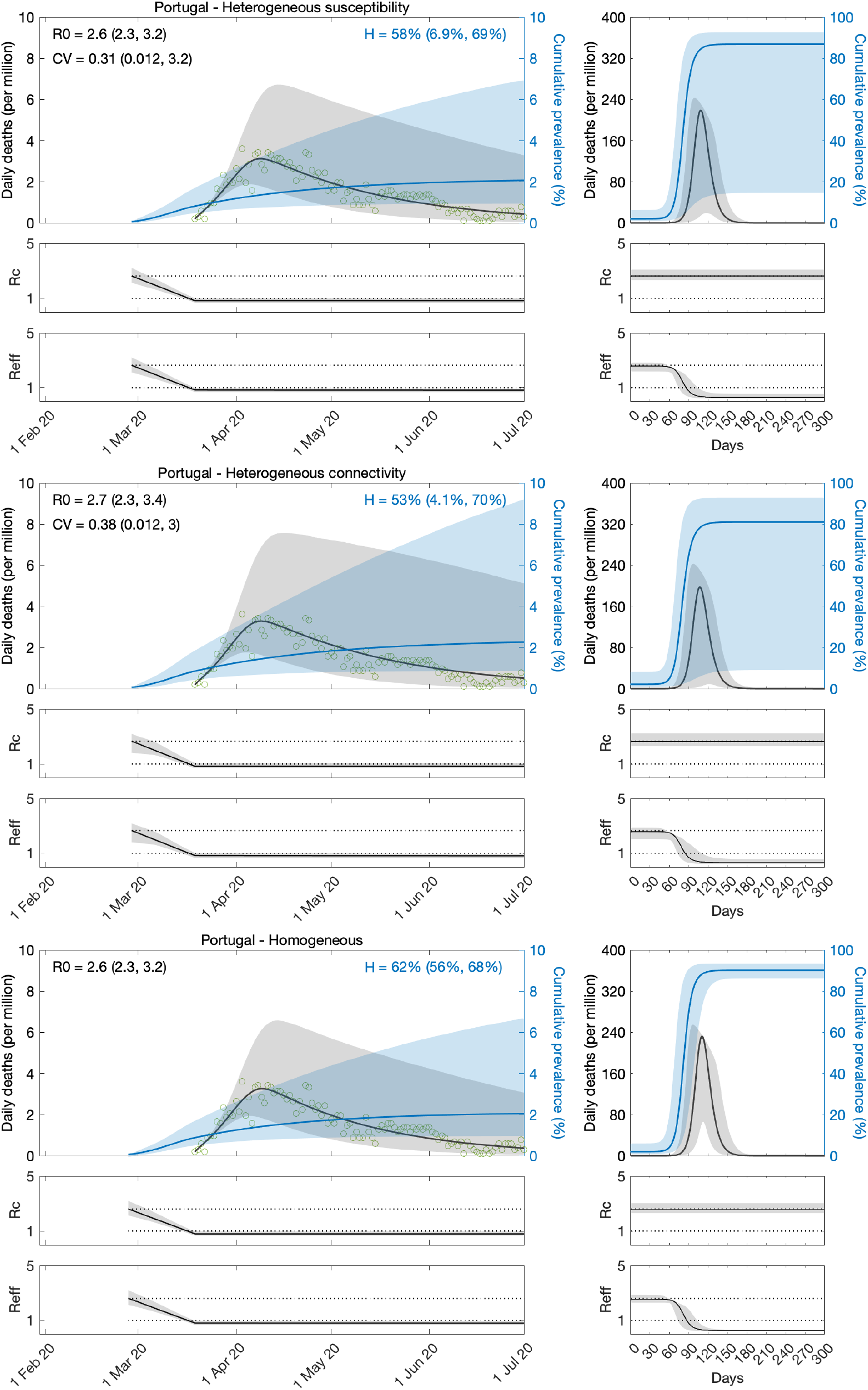
Estimating SARS-CoV-2 transmission in Portugal by fitting one wave of COVID-19 deaths. Variation in susceptibility (top panels); variation in connectivity (middle panels); and homogeneous model (bottom panels). Susceptibility or connectivity factors implemented as gamma distributions. Controlled (*ℛ*_*c*_) and effective (*ℛ*_*eff*_) reproduction numbers are displayed on shallow panels underneath the main plots. Basic reproduction number, coefficients of variation and transmissibility profile parameters estimated by Bayesian inference as described in Methods (estimates in Table 2). Curves represent model reconstructions from the median posterior parameter estimates. Shades represent 95% credible intervals from 100,000 posterior samples.

We estimate the basic reproduction number *ℛ*_0_ with 95% credible intervals (CI) around 3.5 − 3.8 in Spain and 2.3 − 3.4 in Portugal. For the minimal transmissibility factor *c*_1_ we estimate 0.21 − 0.27 when individual variation is allowed and 0.18 − 0.19 with the homogeneity constraint in Spain, while in Portugal we estimate the wider intervals 0.26 − 0.39. For coefficients of variation in Spain, we estimate CV in the range 1.3 − 2.4 under heterogeneous susceptibility and 1.0 − 1.7 under heterogeneous connectivity. In Portugal, we obtain the much wider and uninformative ranges 0.0 − 3.2.

Left plots in Figures 3 and 4 show the best fitting model solutions generated from the median posterior estimates of each parameter in the respective countries as well as the 95% CI generated from 100,000 posterior samples. The herd immunity thresholds *ℋ*, calculated from *ℛ*_0_ and CV estimates, are *ℋ* = 19% (95% CI, 13-32%) under heterogeneous susceptibility, *ℋ* = 19% (95% CI, 13-36%) under heterogeneous connectivity, and *ℋ* = 72% (95% CI, 71-73%) when homogeneity is imposed, in Spain. In Portugal, we obtain *ℋ* = 19% (95% CI, 5-69%) under heterogeneous susceptibility, *ℋ* = 13% (95% CI, 3-69%) under heterogeneous connectivity, and *ℋ* = 67% (95% CI, 66-69%) when homogeneity is imposed. Credible intervals for *ℋ* in Portugal are wide and uninformative when individual variation is allowed which is expected given the wide ranges obtained for CV. NPIs in Portugal were initiated very early in the epidemic which resulted in transmissibility reductions blending with *ℛ*_0_, making parameter identification a major challenge from the data accessible to us.

In Spain, where the three models provide good fits to the data, model selection criteria such as AIC (Akaike information criterion) support the heterogeneous implementations. To better distinguish the various models, we run the respective systems of equations forward, under a set of conventions, and compare the respective projected outcomes. Right plots in Figures 3 were generated by taking the end conditions of the left plots, moving all exposed and infectious individuals to recovered (except a residual proportion to seed a new outbreak) and running each model with the estimated *ℛ*_0_ and CV until the susceptible pool has been effectively depleted in all implementations. In this manner we can visualise how much more burden of infection appears to be ahead when models are constrained to be homogeneous (a manifestation of their relatively higher *ℋ*). Roughly, epidemics peak one order of magnitude higher when models are homogeneous. This must have broad implications, which we believe remain largely unappreciated, for how a population will experience an epidemic, irrespective of how this basic scenario is adapted to specific factors such as behavioural patterns, seasonality, viral evolution, or vaccination.

The same analysis applied to Portugal, selects in favour of the homogeneity assumption and larger projected waves. We recall, however, the great uncertainty associated with these specific results. We investigate this further through fittings to longer data series, in the first instance.

### Fitting models to two serial pandemic waves

Here we take the models with heterogeneity in susceptibility (Equations 8-11) and connectivity (Equations 15-18) and apply the transmissibility profile in (Equations 20-22) to both before fitting the model outputs to COVID-19 deaths recorded daily until 1 March 2021, in Spain and Portugal. As before a homogeneous version obtained by setting *ν* = 0 was also fitted. Results for Spain are shown in Table 3 and Figure 5, and for Portugal in Table 4 and Figure 6.

**Table 3:** Model parameters for Spain (two waves). Estimated by Bayesian inference based on daily deaths until 1 March 2021. Model selection based on maximum log-likelihood (LL) and Akaike information criterion (AIC). Best fitting models have lower AIC scores (best in red, second best in blue). Herd immunity threshold (*ℋ*) derived from estimated *ℛ*_0_ and CV (*ν*).

|  | Heterogeneous susceptibility |  | Heterogeneous connectivity |  | Homogeneous |  |
| --- | --- | --- | --- | --- | --- | --- |
|  | Median | 95% CI | Median | 95% CI | Median | 95% CI |
| *Estimated parameters* |  |  |  |  |  |  |
| $T_0$ | 3.22 | [3.18, 3.31] | 6.06 | [6.06, 6.07] | 5.99 | [5.98, 5.99] |
| $T_2$ | 448.42 | [446.58, 452.53] | 429.91 | [427.04, 433.98] | 783.88 | [777.04, 784.12] |
| $c_1$ | 0.20 | [0.20, 0.20] | 0.21 | [0.21, 0.21] | 0.17 | [0.17, 0.17] |
| $c_2$ | 0.71 | [0.71, 0.71] | 0.68 | [0.68, 0.68] | 0.50 | [0.50, 0.50] |
| $\eta$ | 12* | [12.42, 12.50] | 12* | [11.54, 12.45] | 10* | [10.04, 10.07] |
| $\mathcal{R}_0$ | 4.36 | [4.32, 4.37] | 3.93 | [3.92, 3.95] | 4.11 | [4.10, 4.11] |
| $\nu$ | 2.02 | [2.01, 2.02] | 1.30 | [1.28, 1.31] | 0 | - |
| *Derived from estimated parameters* |  |  |  |  |  |  |
| $\mathcal{H}$ | 25% | [25%, 25%] | 27% | [27%, 27 %] | 76% | [76%, 76%] |
| *Model selection* |  |  |  |  |  |  |
| LL | -3511.40 |  | -3269.90 |  | -5826.40 |  |
| AIC | **7036.80** |  | **6553.90** |  | **11665.00** |  |

\* Rounded median resulting in integer number of days.

**Table 4:** Model parameters for Portugal (two waves). Estimated by Bayesian inference based on daily deaths until 1 March 2021. Model selection based on maximum log-likelihood (LL) and Akaike information criterion (AIC). Best fitting models have lower AIC scores (best in red, second best in blue). Herd immunity threshold (*ℋ*) derived from estimated *ℛ*_0_ and CV (*ν*).

|  | Heterogeneous susceptibility |  | Heterogeneous connectivity |  | Homogeneous |  |
| --- | --- | --- | --- | --- | --- | --- |
|  | Median | 95% CI | Median | 95% CI | Median | 95% CI |
| *Estimated parameters* |  |  |  |  |  |  |
| $T_0$ | 0.03 | [0.01, 0.05] | 0.04 | [0.00, 0.10] | 0.05 | [0.00, 0.10] |
| $T_2$ | 585.14 | [573.66, 672.90] | 644.26 | [615.71, 695.38] | 899.67 | [872.69, 946.07] |
| $c_1$ | 0.27 | [0.27, 0.32] | 0.24 | [0.23, 0.25] | 0.17 | [0.16, 0.18] |
| $c_2$ | 0.71 | [0.71, 0.87] | 0.71 | [0.70, 0.77] | 0.66 | [0.64, 0.68] |
| $c_3$ | 0.34 | [0.27, 0.48] | 0.42 | [0.39, 0.48] | 0.35 | [0.34, 0.36] |
| $\eta$ | 16* | [15.52, 17.39] | 14* | [12.76, 15.34] | 12* | [11.54, 12.33] |
| $\mathcal{R}_0$ | 2.94 | [2.66, 2.96] | 3.25 | [3.14, 3.32] | 4.36 | [4.20, 4.58] |
| $\nu$ | 0.55 | [0.54, 0.59] | 0.47 | [0.47, 0.49] | 0 | - |
| *Derived from estimated parameters* |  |  |  |  |  |  |
| $\mathcal{H}$ | 56% | [52%, 57%] | 56% | [54%, 57%] | 77% | [76%, 78%] |
| *Model selection* |  |  |  |  |  |  |
| LL | -1550.80 |  | -1376.80 |  | -1234.30 |  |
| AIC | **3117.50** |  | **2769.50** |  | **2482.50** |  |

420 \* Rounded median resulting in integer number of days.

**Figure 5:**
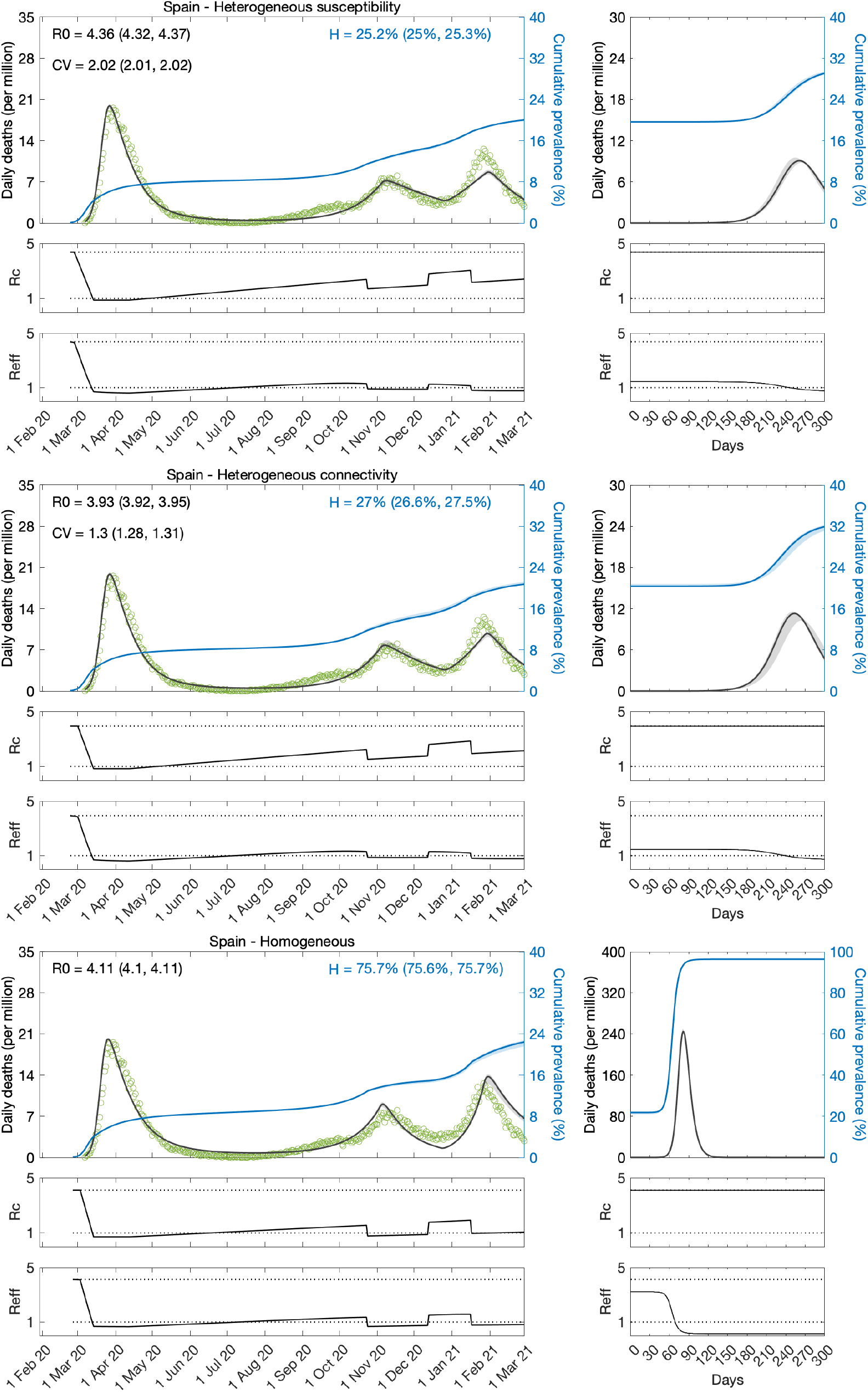
Estimating SARS-CoV-2 transmission in Spain by fitting two serial waves of COVID-19 deaths. Variation in susceptibility (top panels); variation in connectivity (middle panels); and homogeneous model (bottom panels). Susceptibility or connectivity factors implemented as gamma distributions. Controlled (*ℛ*_*c*_) and effective (*ℛ*_*eff*_) reproduction numbers are displayed on shallow panels underneath the main plots. Basic reproduction number, coefficients of variation and transmissibility profile parameters estimated by Bayesian inference as described in Methods (estimates in Table 3). Curves represent model reconstructions from the median posterior parameter estimates. Shades represent 95% credible intervals from 100,000 posterior samples.

**Figure 6:**
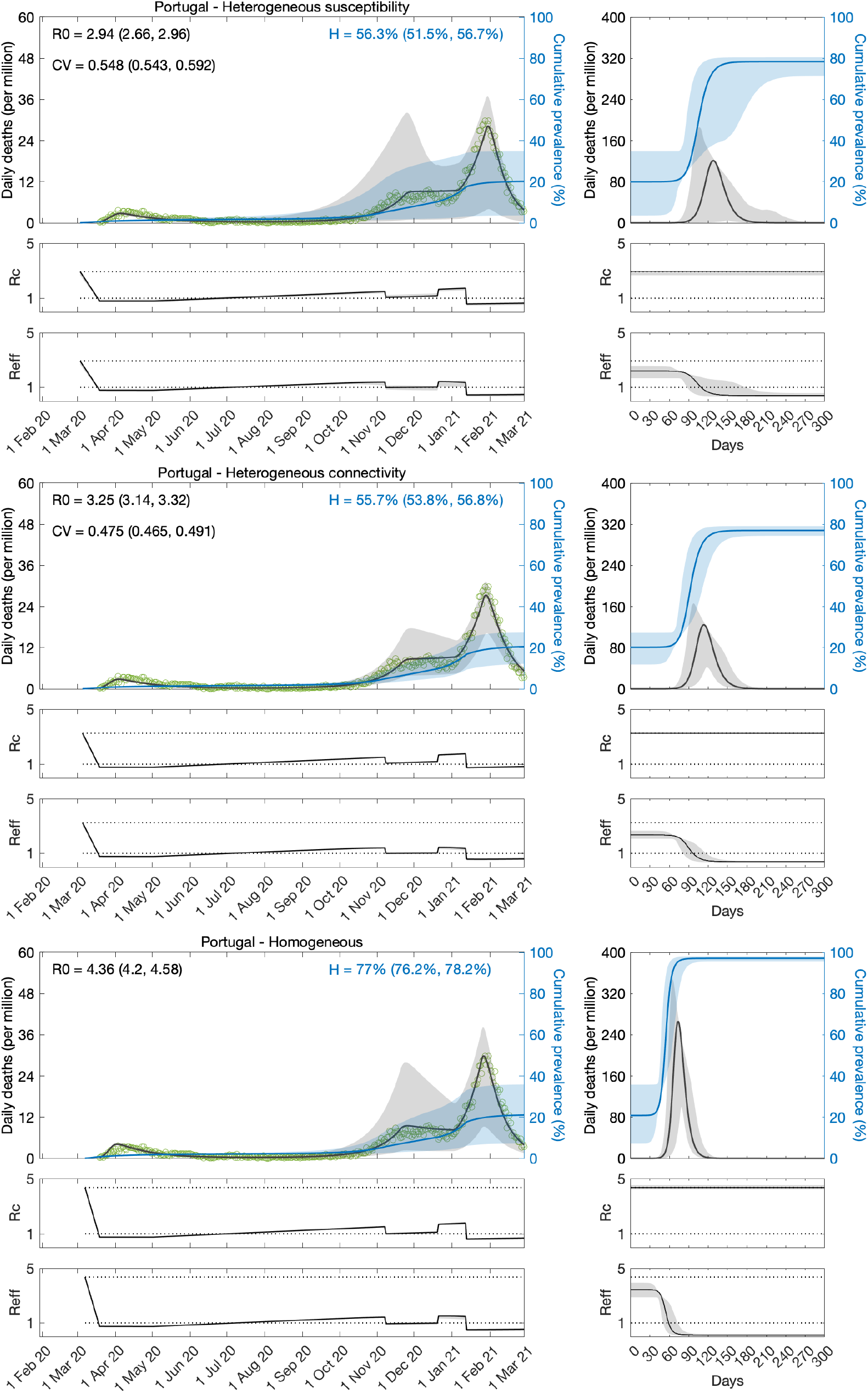
Estimating SARS-CoV-2 transmission in Portugal by fitting two serial waves of COVID-19 deaths. Variation in susceptibility (top panels); variation in connectivity (middle panels); and homogeneous model (bottom panels). Susceptibility or connectivity factors implemented as gamma distributions. Controlled (*ℛ*_*c*_) and effective (*ℛ*_*eff*_) reproduction numbers are displayed on shallow panels underneath the main plots. Basic reproduction number, coefficients of variation and transmissibility profile parameters estimated by Bayesian inference as described in Methods (estimates in Table 4). Curves represent model reconstructions from the median posterior parameter estimates. Shades represent 95% credible intervals from 100,000 posterior samples.

We estimate *ℛ*_0_ with 95% CI around 3.9 − 4.4 in Spain and 2.7 − 4.6 in Portugal. For the minimal transmissibility factor *c*_1_ we estimate 0.20 − 0.21 when individual variation is allowed and 0.17 with the homogeneity constraint in Spain, while in Portugal we obtain 0.23 − 032 with individual variation and 0.16 − 018 without. For coefficients of variation in Spain, we estimate CV around 2.0 under heterogeneous susceptibility and 1.3 under heterogeneous connectivity. In Portugal, the estimates are around 0.5.

Left plots in Figures 5 and 6 show best fitting model solutions as well as the respective 95% CI. In Spain, basic herd immunity thresholds calculated from best fitting *ℛ*_0_ and CV are *ℋ* = 25% under heterogeneous susceptibility, *ℋ* = 27% under heterogeneous connectivity, and *ℋ* = 76% when homogeneity is imposed. In Portugal, we obtain again wider and higher ranges: *ℋ* = 56% (95% CI, 52-57%) under heterogeneous susceptibility, *ℋ* = 56% (95% CI, 54-57%) under heterogeneous connectivity, and *ℋ* = 77% (95% CI, 76-78%) when homogeneity is imposed.

Puzzling, as in the case of shorter data series, model selection supports the homogeneous model for Portugal while still favouring the incorporation of individual variation in Spain. Moreover, for each country, results are consistent whether we base our estimates on one or two waves of the national epidemic. This consistency was also verified in England and Scotland (Gomes et al 2022). This suggests that the earlier initiation of NPIs may not fully explain why results for Portugal contrast with those for other countries studied. In the next section we explore whether this may be due to the asynchrony between the two largest regions (comprising approximately 66% of the total population), which could ultimately result in a mis-specified model for this country.

### Fitting models to regional data in Portugal

Intrigued by the puzzling results for Portugal at country level, we gathered regional data. We found that the epidemic dynamics were considerably different between the two largest regions: North, home to roughly one third of the Portuguese population; and Lisbon and Tagus Valley, home to another third. Asynchrony of epidemic dynamics between regions of similar sizes may require disaggregated analyses. We then decided to fit the mortality data for the two regions simultaneously, estimating common parameters to describe country-wide lockdowns, and regions-specific parameters to describe the basic transmission dynamics (*ℛ*_0_ and CV). The results are provided in Tables 5, 6 and Figures 7-9. According to these analyses, the best-fitting models include heterogeneity.

**Table 5:** Model parameters for the North and Lisbon regions of Portugal (one wave). Estimated by Bayesian inference based on daily deaths until 1 July 2020. Model selection based on maximum log-likelihood (LL) and Akaike information criterion (AIC). Best fitting models have lower AIC scores (best in red, second best in blue). Herd immunity threshold (*ℋ*) derived from estimated *ℛ*_0_ and CV (*ν*).

|  | Heterogeneous susceptibility |  | Heterogeneous connectivity |  | Homogeneous |  |
| --- | --- | --- | --- | --- | --- | --- |
|  | Median | 95% CI | Median | 95% CI | Median | 95% CI |
| *Common parameters* |  |  |  |  |  |  |
| $T_0$ | 0.05 | [0.00, 0.10] | 0.05 | [0.00, 0.10] | 0.05 | [0.00, 0.10] |
| $c_1$ | 0.48 | [0.43, 0.52] | 0.55 | [0.47, 0.61] | 0.43 | [0.40, 0.47] |
| $\eta$ | 24* | [19.45, 28.99] | 21* | [14.18, 27.37] | 29* | [28.50, 28.50] |
| *North* |  |  |  |  |  |  |
| $\mathcal{R}_0$ | 2.17 | [2.00, 2.45] | 2.43 | [2.17, 2.92] | 1.91 | [1.80, 2.09] |
| $v$ | 3.44 | [3.38, 3.46] | 3.45 | [3.39, 3.46] | 0 | - |
| $\mathcal{H}$ | 5.9% | [5.2%, 7.0%] | 3.5% | [3.0%, 4.4%] | 48% | [44%, 52%] |
| *Lisbon and Tagus Valley* |  |  |  |  |  |  |
| $\mathcal{R}_0$ | 1.78 | [1.65, 2.00] | 1.65 | [1.49, 1.96] | 1.85 | [1.74, 2.02] |
| $v$ | 0.07 | [0.01, 0.56] | 0.07 | [0.01, 0.57] | 0 | - |
| $\mathcal{H}$ | 44% | [32%, 50%] | 39% | [21%, 49%] | 46% | [43%, 51%] |
| *Model selection* |  |  |  |  |  |  |
| LL | -520.56 |  | -479.75 |  | -578.50 |  |
| AIC | **1055.10** |  | **973.49** |  | **1167.00** |  |

\* Rounded median resulting in integer number of days.

**Table 6:**
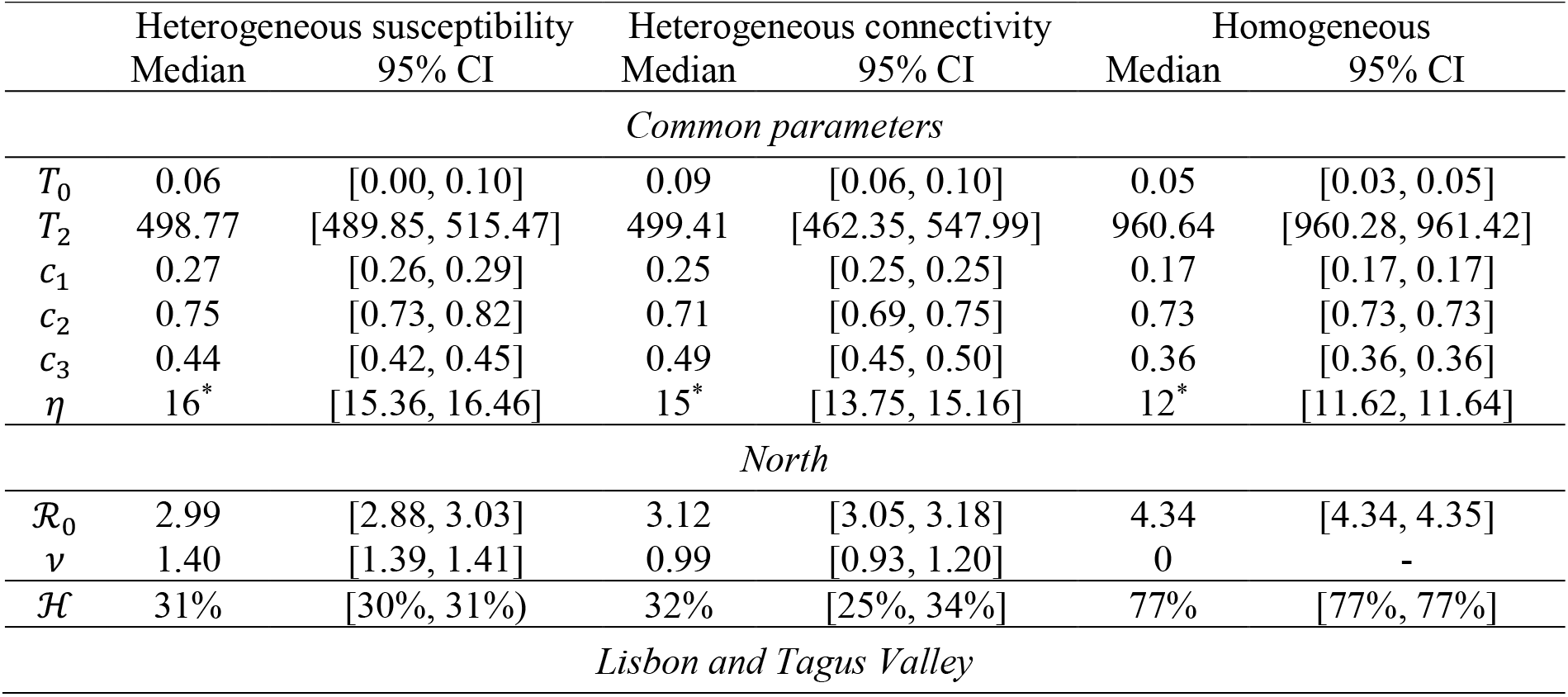
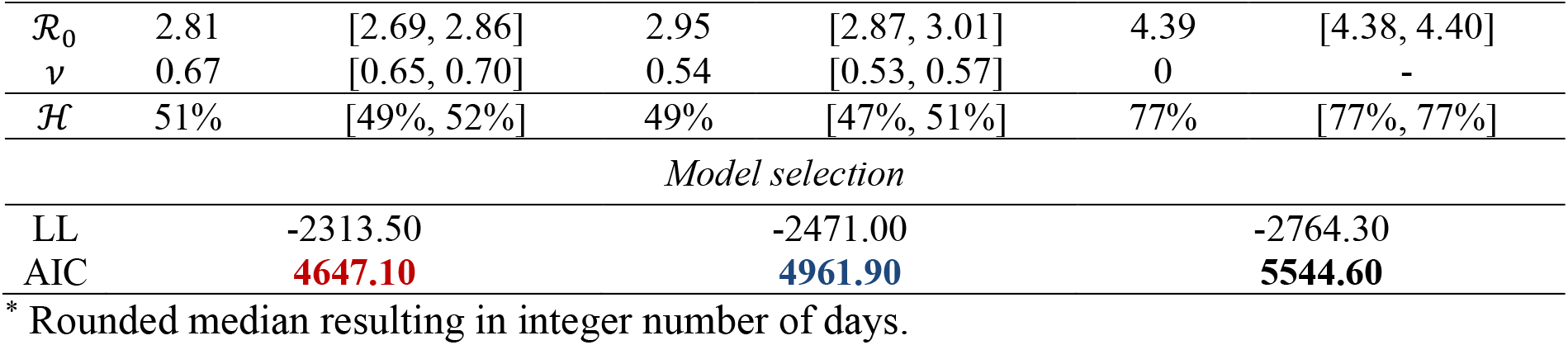
Model parameters for the North and Lisbon regions of Portugal (two waves). Estimated by Bayesian inference based on daily deaths until 1 March 2021. Model selection based on maximum log-likelihood (LL) and Akaike information criterion (AIC). Best fitting models have lower AIC scores (best in red, second best in blue). Herd immunity threshold (*ℋ*) derived from estimated *ℛ*_0_ and CV (*ν*).

|  | Heterogeneous susceptibility |  | Heterogeneous connectivity |  | Homogeneous |  |
| --- | --- | --- | --- | --- | --- | --- |
|  | Median | 95% CI | Median | 95% CI | Median | 95% CI |
| *Common parameters* |  |  |  |  |  |  |
| $T_0$ | 0.06 | [0.00, 0.10] | 0.09 | [0.06, 0.10] | 0.05 | [0.03, 0.05] |
| $T_2$ | 498.77 | [489.85, 515.47] | 499.41 | [462.35, 547.99] | 960.64 | [960.28, 961.42] |
| $c_1$ | 0.27 | [0.26, 0.29] | 0.25 | [0.25, 0.25] | 0.17 | [0.17, 0.17] |
| $c_2$ | 0.75 | [0.73, 0.82] | 0.71 | [0.69, 0.75] | 0.73 | [0.73, 0.73] |
| $c_3$ | 0.44 | [0.42, 0.45] | 0.49 | [0.45, 0.50] | 0.36 | [0.36, 0.36] |
| $\eta$ | 16* | [15.36, 16.46] | 15* | [13.75, 15.16] | 12* | [11.62, 11.64] |
| *North* |  |  |  |  |  |  |
| $\mathcal{R}_0$ | 2.99 | [2.88, 3.03] | 3.12 | [3.05, 3.18] | 4.34 | [4.34, 4.35] |
| $v$ | 1.40 | [1.39, 1.41] | 0.99 | [0.93, 1.20] | 0 | - |
| $\mathcal{H}$ | 31% | [30%, 31%] | 32% | [25%, 34%] | 77% | [77%, 77%] |
| *Lisbon and Tagus Valley* |  |  |  |  |  |  |
| $\mathcal{R}_0$ | 2.81 | [2.69, 2.86] | 2.95 | [2.87, 3.01] | 4.39 | [4.38, 4.40] |
| $\nu$ | 0.67 | [0.65, 0.70] | 0.54 | [0.53, 0.57] | 0 | - |
| $\mathcal{H}$ | 51% | [49%, 52%] | 49% | [47%, 51%] | 77% | [77%, 77%] |

*Model selection*
|  |  |  |  |
| --- | --- | --- | --- |
| LL | -2313.50 | -2471.00 | -2764.30 |
| AIC | **4647.10** | **4961.90** | **5544.60** |

\* Rounded median resulting in integer number of days.

**Figure 7:**
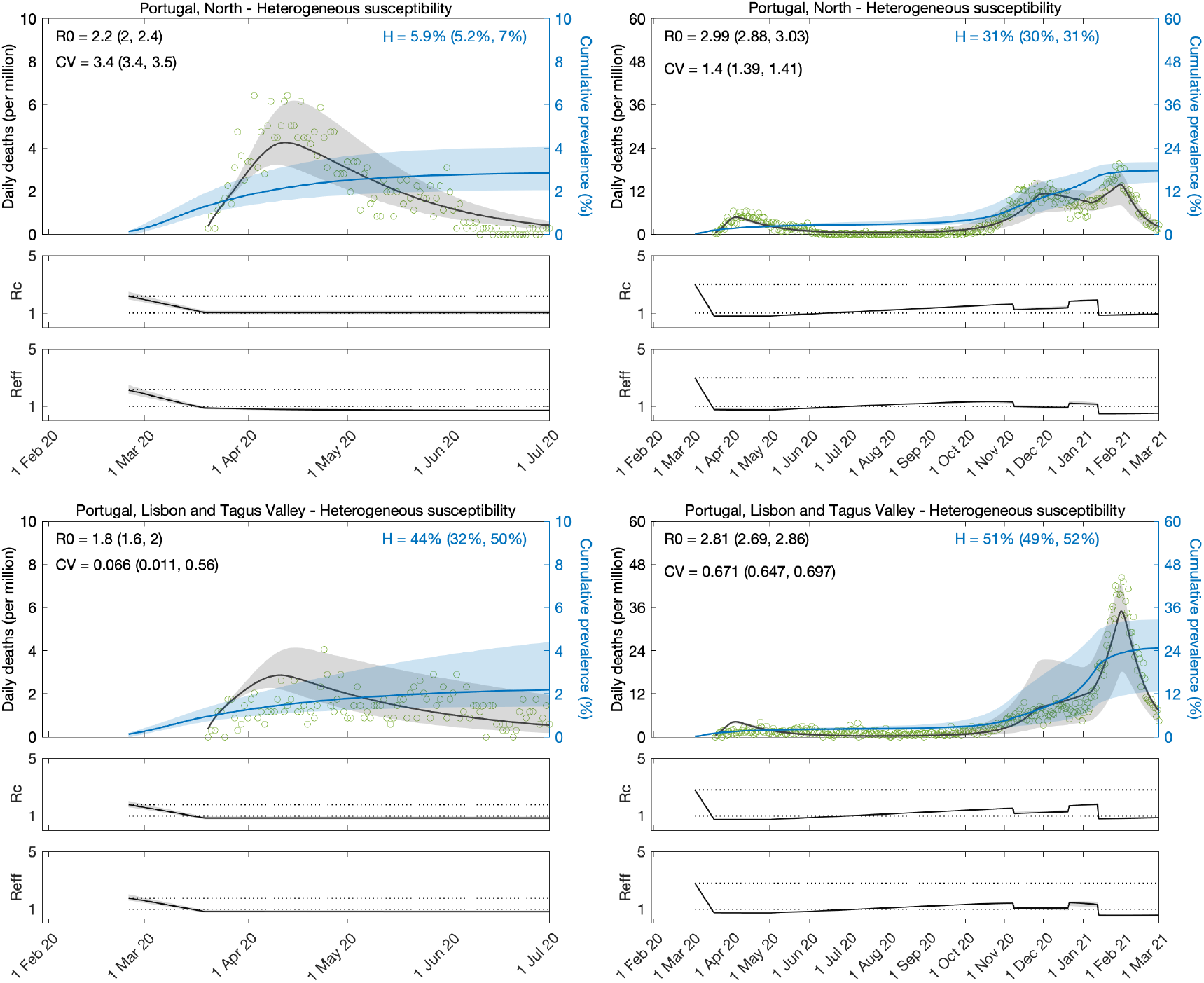
Estimating SARS-CoV-2 transmission in the two larger regions of Portugal. Heterogeneous susceptibility implemented as a gamma distribution. Controlled (*ℛ*_*c*_) and effective (*ℛ*_*eff*_) reproduction numbers are displayed on shallow panels underneath the main plots. Basic reproduction number, coefficients of variation and transmissibility profile parameters estimated by Bayesian inference as described in Methods (estimates in Table 5). Curves represent model reconstructions from the median posterior parameter estimates. Shades represent 95% credible intervals from 100,000 posterior samples.

**Figure 8:**
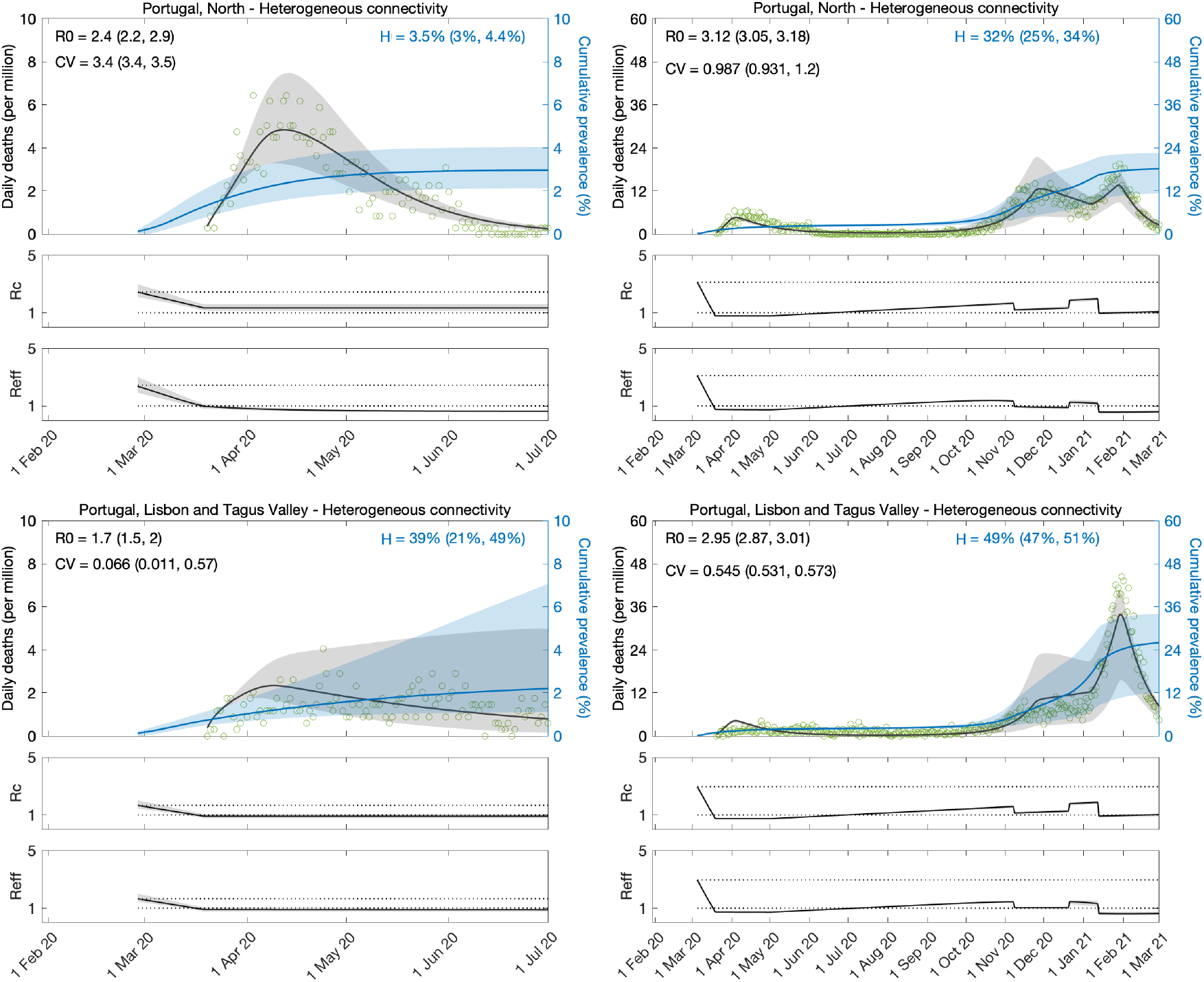
Estimating SARS-CoV-2 transmission in the two larger regions of Portugal. Heterogeneous connectivity implemented as a gamma distribution. Controlled (*ℛ*_*c*_) and effective (*ℛ*_*eff*_) reproduction numbers are displayed on shallow panels underneath the main plots. Basic reproduction number, coefficients of variation and transmissibility profile parameters estimated by Bayesian inference as described in Methods (estimates in Table 5). Curves represent model reconstructions from the median posterior parameter estimates. Shades represent 95% credible intervals from 100,000 posterior samples.

**Figure 9:**
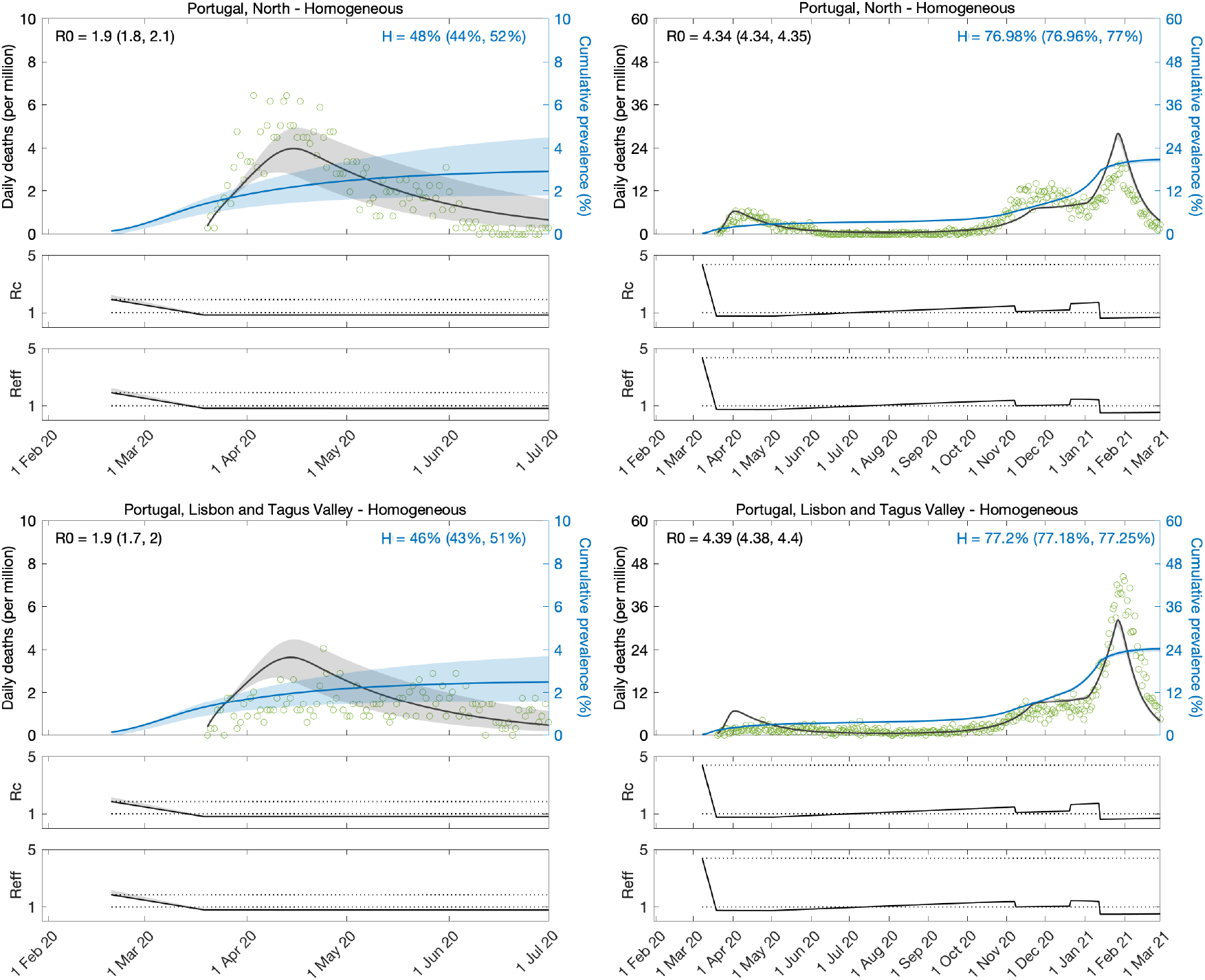
Estimating SARS-CoV-2 transmission in the two larger regions of Portugal. Homogeneous model. Controlled (*ℛ*_*c*_) and effective (*ℛ*_*eff*_) reproduction numbers are displayed on shallow panels underneath the main plots. Basic reproduction number, coefficients of variation and transmissibility profile parameters estimated by Bayesian inference as described in Methods (estimates in Table 5). Curves represent model reconstructions from the median posterior parameter estimates. Shades represent 95% credible intervals from 100,000 posterior samples.

Herd immunity thresholds inferred by fits to the longer series (until 1 March 2021) are around 25 − 34% in the North (similar as Spain, England and Scotland) and 47 − 52% in Lisbon and Tagus Valley. The higher *ℋ* in the capital region results from the estimation of a lower CV. This may be real and due to the more urban character of Lisbon and Tagus Valley, or, in contrast, it may be a spurious result of the lack of a first wave in the region to inform the model. It would be interesting to replicate the regional analysis in other countries to investigate to what extent more urban regions have higher HIT. As for the North, *ℋ* is much closer to that of Spain, England and Scotland, but slightly higher, nevertheless. This may also be a slightly spurious consequence of the first wave being more suppressed there (although not as much as in Lisbon and Tagus Valley) than in those other nations. Alternatively, there may be differences in reporting across countries (particularly those affecting whether deaths are declared with or from COVID-19 [Ferreira 2022; Gonçalves 2022]) influencing the accuracy of estimated model parameters.

Fits to the shorter series (until 1 July 2020) are again inconclusive. Not only the uncertainty around parameter estimates is large but some estimates are implausible. First, *ℛ*_0_ around 2 or less is on the low end of consensus estimates (Flaxman et al. 2020; Keeling et al. 2020; Viana et al. 2021; Wood 2021). More strikingly, the algorithm is incapable of estimating CV from these regional series, resulting in convergence to the upper and lower limits of the prior distribution (uniform between 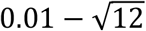) in the North and Lisbon regions, respectively.

In summary, the results reported in this section support two notions. First, asynchronous dynamics may compromise parameter estimation based on model fittings to aggregated data. This should depend on whether the asynchrony in question is between similar-sized regions. Second, the early estimation of parameters for heterogeneous models (especially CV) may require a larger first wave than that required by models that are either homogeneous (Flaxman et al. 2020; Wood 2021) or have their heterogeneity informed directly by specific data (Keeling et al. 2020). The downside of these other approaches, however, is that heterogeneity is either absent or possibly incomplete, resulting in reduced selection and biased estimates. Among the studies we have completed, England, Scotland and Spain had sufficiently sized first waves to inform the inference of CV while Portugal did not.

## Discussion

We fitted SEIR models, with inbuilt distributions of individual susceptibility or exposure to infection, to daily series of COVID-19 deaths in Spain and Portugal. We estimated relevant transmission parameters, such as the basic reproduction number *ℛ*_0_, and a time dependent transmissibility profile *c*(*t*) which multiplies *ℛ*_0_ to account for effects of NPIs, seasonality, viral evolution, or any unexplicit factor that modifies the ability of the virus to infect new hosts. In addition, as in (Gomes et al. 2022) we estimated coefficients of variation that characterise distributions of unmeasured individual susceptibility or connectivity.

The inference of selectable variation as presented here is uncommon in infectious disease modelling. Prior to attempting this in the context of the COVID-19 pandemic, we and others have conducted related studies in systems that were either experimentally controlled (Dwyer et al. 1997; Ben-Ami et al. 2008; Zwart et al. 2011; Pessoa et al. 2014; Langwig et al. 2017; King et al. 2018) or already endemic (Smith et al. 2005; Bellan et al. 2015; Corder et al. 2020). Several aspects of the pandemic made it more challenging. There was an urgency for early results from inherently scarce data, a challenge amplified by non-pharmaceutical interventions designed to suppress the epidemic. In countries where interventions began earlier (relative to the epidemic momentum), such as Portugal, it has been impossible to reach conclusive results without finer data and methods. In Spain (this study) and England and Scotland (Gomes et al 2022), on the other hand, interventions started later, and results were consistent, both internally and between each other. Our coefficients of variation for individual connectivity are similar to those measured directly by contact surveys (Gomes et al 2022).

Another group of authors highlighted the importance of considering the interplay between social dynamics and spread of infection when interpreting coefficients of variation (Tkachenko 2021). If society changed over time in such a way that individuals with low susceptibility/exposure in one wave became high susceptibility/exposure in a later wave, then coefficients of variation estimated from two-wave fits should be lower than those estimated from one-wave fits, resulting in higher herd immunity thresholds. Our results for Spain are not indicative of this process playing a significant role in COVID-19. We estimate CV around 1.41 (95%, 0.97 − 1.74) when we fit the first wave only, and 1.30 (95%, 1.28 − 1.31) when we fit two waves. The two-wave estimate is not significantly lower than that obtained from the one-wave analysis, suggesting that the postulated mechanism is not affecting our inferences. We saw the same consistency in our previous analysis of England and Scotland (Gomes et al. 2022). We find this unsurprising given the amply reported evidence of socioeconomic determinants as key drivers of heterogeneity in infectious diseases (e.g., Millett et al. 2020, Xia et al. 2022). Societal changes may not significantly impact our inferences unless major inversions in socioeconomic gradients had occurred which is unimaginable in the time scale of a pandemic. On the contrary, the opposite seems more plausible as more disadvantaged social groups suffer more from both disease and containment measures, exacerbating preexisting heterogeneity (Okonkwo et al. 2021).

The exploration presented here for Spain confirms recent findings for England and Scotland that the original SARS-CoV-2 had a herd immunity threshold in the range 20-30%. The main specificity of the underlying studies is to include individual variation in susceptibility and exposure to infection in the set of model parameters being estimated. The modelling approach is relatively new, and its limits of applicability remain a subject for research and further methodological developments. Here we adopt a dataset from Portugal to highlight features that compromise the applicability of the basic method and, in some instances, propose refinements to push those limits.

## Data Availability

Datasets are publicly available at the respective national ministry of health websites.

## Acknowledgements

We thank Rodrigo Corder, Jessica King and Antonio Montalbán for technical discussions and contributions to related research.

## Author contributions

M.G.M.G. conceived the study. R.A. and M.G.M.G. performed the analyses. All authors interpreted the data and wrote the paper.

## Data availability

We used publicly available data from the coronavirus dashboards for Spain [https://cnecovid.isciii.es/covid19] and Portugal [https://covid19.min-saude.pt/ponto-de-situacao-atual-em-portugal]. Population sizes were obtained from recent censuses: 46,771,836 for Spain; 10,196,709 for Portugal; 3,573,000 for Portugal-North; and 3,447,173 for Lisbon and Tagus Valley.

