## Supplementary information for "Herd immunity thresholds for SARS-CoV-2 estimated from unfolding epidemics"

### Supplementary Information for "Herd immunity thresholds for SARS-CoV-2 estimated from unfolding epidemics": Exploratory sensitivity analyses

Ricardo Aguas[1], Guilherme Gonçalves[2], Marcelo U. Ferreira[3], M. Gabriela M. Gomes[4,5]*

[1] *Centre for Tropical Medicine and Global Health, Nuffield Department of Medicine, University of Oxford, Oxford, United Kingdom.*

[2] *Unidade Multidisciplinar de Investigação Biomédica, Instituto de Ciências Biomédicas Abel Salazar, Universidade do Porto, Porto, Portugal.*

[3] *Instituto de Ciências Biomédicas, Universidade de São Paulo, São Paulo, Brazil.*

[4] *Department of Mathematics and Statistics, University of Strathclyde, Glasgow, United Kingdom.*

[5] *Centro de Matemática e Aplicações, Faculdade de Ciências e Tecnologia, Universidade Nova de Lisboa, Caparica, Portugal.*

#### Contents:

1. Sensitivity to random mixing.

#### 1. Sensitivity to random mixing

In the main text we assumed random mixing among individuals, but human connectivity patterns may be assortative due to societal structures and human behaviours. To explore the sensitivity of our results to deviations from random mixing, we develop an extended formalism that allows individuals to connect preferentially with those with similar connectivity, formally:

$$\lambda(x) = \frac{\beta}{N}\frac{\int y\, h(y-x)[\rho E(y) + I(y)]\, dy}{\int y g(y)\, dy}, \tag{S1}$$

where $h(y-x)$ is a normal distribution on the difference between connectivity factors (Figure S1).

In this case we write the effective reproduction number more generally as the incidence of new infections divided by the total number of active infections (affected by $\rho$ for individuals in $E$) multiplied by the average duration of infection (also affected by $\rho$ for individuals in $E$)

$$\mathcal{R}_{eff}(t) = \frac{\int \lambda(x,t) x[S(x,t) + \sigma R(x,t)]\, dx}{\int \rho E(x,t) + I(x,t)\, dx}\left(\frac{\rho}{\delta} + \frac{1}{\gamma}\right). \tag{S2}$$

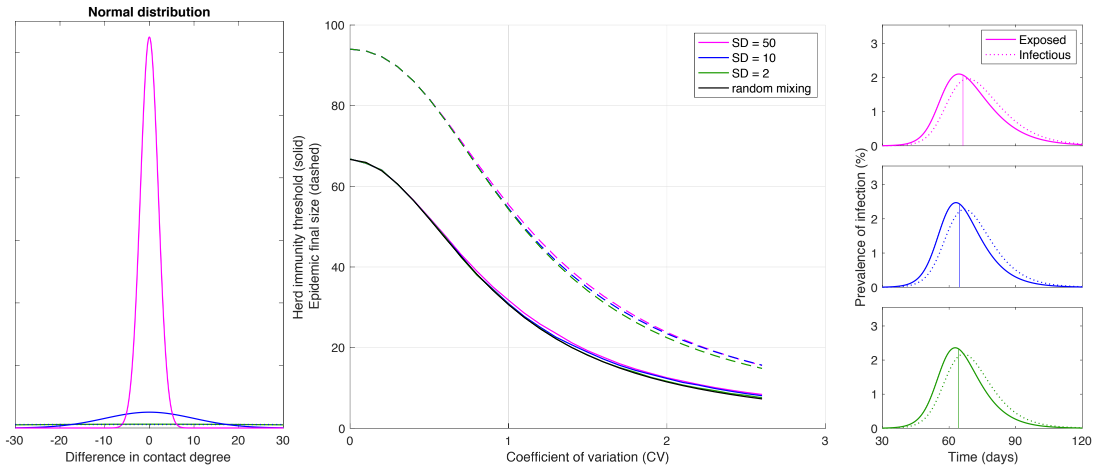


**Figure S1: Herd immunity threshold and epidemic final size with gamma-distributed exposure to infection and assortative mixing.** Curves in central panel generated with the SEIR model (Equation 1-4) assuming $\mathcal{R}_0 = 3$ and gamma-distributed connectivity. Assortative mixing is implemented by imposing a normal distribution for contact preferences such that individuals contact preferentially with those with the similar contact degree (left). This illustration used normal distributions with standard deviation $SD = 50$ (green); $SD = 10$ (blue); and $SD = 2$ (magenta). More assortative mixing leads to more skewed epidemics. Herd immunity thresholds were calculated numerically as the percentage of the population no longer susceptible when new outbreaks are effectively prevented (approximately when the exposed fraction crosses the peak in the absence of mitigation). Final sizes of the corresponding unmitigated epidemics are also shown. Representative epidemics are depicted on the right based on the rightmost CVs represented on the main panel (with vertical lines marking the point when herd immunity is achieved).
